# Performance of DeepSeek, Qwen 2.5 MAX, and ChatGPT Assisting in Diagnosis of Corneal Eye Diseases, Glaucoma, and Neuro-Ophthalmology Diseases Based on Clinical Case Reports

**DOI:** 10.1101/2025.03.14.25323836

**Authors:** Zain S. Hussain, Mohammad Delsoz, Muhammad Elahi, Brian Jerkins, Elliot Kanner, Claire Wright, Wuqaas M. Munir, Mohammad Soleimani, Ali Djalilian, Priscilla A. Lao, Joseph W. Fong, Malik Y. Kahook, Siamak Yousefi

**Author notes:** Correspondence to: Siamak Yousefi, 930 Madison Ave., Suite 471, Memphis, TN 38163, USA, 21, 22 Conflict of Interest: Mohammad Delsoz, Amr Hassan, Amin Nabavi, Amir Rahdar, Brian Fowler, 23 Natalie C. Kerr, Lauren Claire Ditta, Mary E. Hoehn, Margaret M DeAngelis, Andrzej Grzybowski, 24 and Yih-Chung Tham have nothing to disclose. Siamak Yousefi: Received prototype instruments 25 from Remidio, M&S Technologies, and Visrtucal Fields. He gives consultations to the InsihgtAEye 26 and Enolink. signifies first co-authorship and equivalent work performed (Zain S. Hussain, Mohammad Delsoz).

## Abstract

**Background:** This study evaluates the diagnostic performance of several AI models, including Deepseek, in diagnosing corneal diseases, glaucoma, and neuro□ophthalmologic disorders.

**Methods:** We retrospectively selected 53 case reports from the Department of Ophthalmology and Visual Sciences at the University of Iowa, comprising 20 corneal disease cases, 11 glaucoma cases, and 22 neuro□ophthalmology cases. The case descriptions were input into DeepSeek, ChatGPT□4.0, ChatGPT□01, and Qwens 2.5 Max. These responses were compared with diagnoses rendered by human experts (corneal specialists, glaucoma attendings, and neuro□ophthalmologists). Diagnostic accuracy and interobserver agreement, defined as the percentage difference between each AI model’s performance and the average human expert performance, were determined.

**Results:** DeepSeek achieved an overall diagnostic accuracy of 79.2%, with specialty-specific accuracies of 90.0% in corneal diseases, 54.5% in glaucoma, and 81.8% in neuro□ophthalmology. ChatGPT□01 outperformed the other models with an overall accuracy of 84.9% (85.0% in corneal diseases, 63.6% in glaucoma, and 95.5% in neuro□ophthalmology), while Qwens exhibited a lower overall accuracy of 64.2% (55.0% in corneal diseases, 54.5% in glaucoma, and 77.3% in neuro□ophthalmology). Interobserver agreement analysis revealed that in corneal diseases, DeepSeek differed by –3.3% (90.0% vs 93.3%), ChatGPT□01 by –8.3%, and Qwens by –38.3%. In glaucoma, DeepSeek outperformed the human expert average by +3.0% (54.5% vs 51.5%), while ChatGPT□4.0 and ChatGPT□01 exceeded it by +12.1%, and Qwens was +3.0% above the human average. In neuro□ophthalmology, DeepSeek and ChatGPT□4.0 were 9.1% lower than the human average, ChatGPT□01 exceeded it by +4.6%, and Qwens was 13.6% lower.

**Conclusions:** ChatGPT□01 demonstrated the highest overall diagnostic accuracy, especially in neuro□ophthalmology, while DeepSeek and ChatGPT□4.0 showed comparable performance. Qwens underperformed relative to the other models, especially in corneal diseases. Although these AI models exhibit promising diagnostic capabilities, they currently lag behind human experts in certain areas, underscoring the need for a collaborative integration of clinical judgment.

**Plain Language Summary:** This study evaluated how well several artificial intelligence (AI) models diagnose eye diseases compared to human experts. We tested four AI systems across three types of eye conditions: diseases of the cornea, glaucoma, and neuro-ophthalmologic disorders. Overall, one AI model, ChatGPT-01, performed the best, correctly diagnosing about 85% of cases, and it excelled in neuro-ophthalmology by correctly diagnosing 95.5% of cases. Two other models, DeepSeek and ChatGPT-4.0, each achieved an overall accuracy of around 79%, while the Qwens model performed lower, with an overall accuracy of about 64%. When compared with human experts, who achieved very high accuracy in corneal diseases (93.3%) and neuro-ophthalmology (90.9%) but lower in glaucoma (51.5%), the AI models showed mixed results. In glaucoma, for instance, some AI models even outperformed human experts slightly, while in corneal diseases, all AI models were less accurate than the experts. These findings indicate that while AI shows promise as a supportive tool in diagnosing eye conditions, it still needs further improvement. Combining AI with human clinical judgment appears to be the best approach for accurate eye disease diagnosis.

**Key summary points:**

- **Why carry out this study?** With the rising burden of eye diseases and the inherent diagnostic challenges for complex conditions like glaucoma and neuro-ophthalmologic disorders, there is an unmet need for innovative diagnostic tools to support clinical decision-making.
- **What did the study ask?** This study evaluated the diagnostic performance of four AI models across three ophthalmologic subspecialties, testing the hypothesis that advanced language models can achieve accuracy levels comparable to human experts.
- **What was learned from the study?** Our results showed that ChatGPT-01 achieved the highest overall accuracy (84.9%), excelling in neuro-ophthalmology with a 95.5% accuracy, while DeepSeek and ChatGPT-4.0 each achieved 79.2%, and Qwens reached 64.2%.
- **What specific outcomes were observed?** In glaucoma, AI model accuracies ranged from 54.5% to 63.6%, with some models slightly surpassing the human expert average of 51.5%, underscoring the diagnostic difficulty of this condition.
- **What has been learned and future implications?** These findings highlight the potential of AI as a valuable adjunct to clinical judgment in ophthalmology, although further research and the integration of multimodal data are essential to optimize these tools for routine clinical practice.

## Introduction

Glaucoma, corneal diseases, and neuro-ophthalmologic disorders present significant challenges in ophthalmic diagnostics due to their complex clinical presentations and the need for specialized expertise. Glaucoma remains a leading cause of irreversible blindness worldwide^1^, with its early diagnosis complicated by physiological traits like varying optic disc sizes, and certain pathological signs such as non-glaucomatous optic nerve diseases. These can be misleading and distinguishing between different conditions can be challenging^2,3^. Similarly, corneal diseases, which encompass a wide spectrum of infections, dystrophies, degenerations, and traumatic injuries, require timely and precise diagnosis to prevent vision loss. In neuro-ophthalmology, the intricate interplay between neurological and ophthalmic conditions demands a high level of diagnostic acumen, yet the global shortage of neuro-ophthalmologists results in prolonged wait times and delayed treatment^4^. Despite the availability of multiple diagnostic tests, the subjective nature of clinical evaluation leads to considerable variability among clinicians.

The rapid advancement of artificial intelligence (AI) has introduced novel approaches for enhancing medical diagnostics, particularly in ophthalmology. Since the advent of deep convolutional neural networks (CNNs), AI has demonstrated effectiveness in detecting and classifying ocular diseases^5,6^. More recently, large language models (LLMs) have garnered attention for their potential in clinical decision-making^7^. ChatGPT, developed by OpenAI, has exhibited promising capabilities in responding to ophthalmic inquiries and assisting in diagnostic reasoning^8,9^. Previous research has explored ChatGPT’s performance assisting in diagnosing glaucoma, corneal diseases, and neuro-ophthalmic disorders, and other ophthalmic conditions highlighting both its potential and limitations^10-13^. However, the landscape of AI-driven diagnostics continues to evolve with the emergence of new models.

Recently, Chinese-developed AI chatbots such as DeepSeek-V3 (release latest version Dec 2024) and Alibaba’s AI model Qwen 2.5 Max (release latest version Jan 2025) have gained attention^14,15^, yet their capabilities in clinical diagnosis in ophthalmology remain unexplored. There are currently no dedicated research papers evaluating their effectiveness compared to ChatGPT or their potential role in ophthalmic medical applications. This study aims to assess the performance of these three advanced LLMs DeepSeek, Qwen 2.5 Max, and ChatGPT in diagnosing glaucoma, corneal diseases, and neuro-ophthalmologic disorders based on detailed clinical case reports. By comparing the diagnostic accuracy of these models with expert ophthalmologists, we seek to evaluate their capabilities, limitations, and potential applications in clinical practice. Understanding the strengths and weaknesses of these AI systems can contribute to the development of more reliable and efficient diagnostic tools, ultimately enhancing patient care and optimizing the allocation of specialized ophthalmic resources.

## Methods

### Data Source and Case Selection

We utilized 53 clinical case reports from the available database provided by the Department of Ophthalmology and Visual Sciences at the University of Iowa (https://webeye.ophth.uiowa.edu/eyeforum/cases.htm). This database contains clinical cases reports categorized by ophthalmic subspecialties. For this study, we selected cases from three major categories: glaucoma, cornea, and neuro-ophthalmology. Each clinical case covered patients background, including their demographics, main symptoms, medical and ocular history, details of presenting illness, chief complaint, and key findings of examination.

#### 1. Glaucoma

We identified 11 cases representing a diverse spectrum of glaucoma, including primary and secondary forms such as open-angle glaucoma, angle-closure glaucoma, neovascular glaucoma, inflammatory glaucoma, and other variants. These cases were chosen to reflect the heterogeneity of glaucoma presentations, from common phenotypes to rare variants.

#### 2. Corneal eye diseases

A total of 20 cases were selected, covering a wide range of corneal pathologies such as infections (e.g., viral and amoebic keratitis), dystrophies (e.g., Fuchs’ endothelial dystrophy), degenerations (e.g., Salzmann’s nodular degeneration), and systemic or drug-induced conditions (e.g., cystinosis and amiodarone-induced deposits).

#### 3. Neuro-Ophthalmology Diseases

We selected 22 cases encompassing a variety of neuro-ophthalmic conditions, including optic nerve disorders (e.g., optic neuritis and traumatic optic neuropathy), cranial nerve palsies (e.g., trochlear nerve palsy), inflammatory and demyelinating diseases (e.g., Acute Disseminated Encephalomyelitis and idiopathic orbital myositis), and structural or syndromic conditions (e.g., neurofibromatosis and thyroid eye disease). These cases were chosen to represent the complexity and diversity of neuro-ophthalmic diseases.

Since the dataset used in this study was publicly accessible and contained no identifiable patient information, approval from an institutional review board (IRB) was not necessary. All diagnoses evaluated are observed in Supplementary Tables 1, 2, and 3.

##### AI Model Selection and Evaluation

The selected cases were input into three AI models including DeepSeek-V3, Alibaba’s Qwen 2.5 Max, ChatGPT-4o, and ChatGPT-o1 to evaluate their diagnostic performance. Each model was provided with the same case details, including patient demographics, history, chief complaint, and examination findings. Cases requiring specialized diagnostic maneuvers or those with overly obvious diagnoses were excluded for all to ensure a balanced evaluation. The models were tasked with identifying the most likely diagnosis for each case. Each LLM was assigned to analyze and respond to the cases independently. To ensure fairness and prevent any bias, we created separate chat instances for each trial. This approach minimized the influence of the models’ Reinforcement Learning from Human Feedback (RLHF) capabilities and their long-term memory features^16,17^, allowing for an unbiased evaluation of their diagnostic performance. Their responses were evaluated based on diagnostic accuracy and compared with human experts. Interobserver agreements were calculated.

##### Statistical Analysis

Diagnostic accuracy was calculated as the percentage of correct diagnoses relative to the total number of cases evaluated for each ophthalmologic specialty and overall. Interobserver agreement was quantified as the absolute difference in percentage accuracy between each AI model and the average human expert performance.

## Results

A comprehensive evaluation was conducted across 53 cases spanning three ophthalmologic specialties—20 cases in corneal diseases, 11 in glaucoma, and 22 in neuro□ophthalmology—to assess the diagnostic performance of several AI models. DeepSeek correctly diagnosed 90% of corneal disease cases (18/20), achieved an accuracy of 54.5% in glaucoma (6/11), and reached 81.8% accuracy in neuro□ophthalmology (18/22), resulting in an overall accuracy of 79.2% (42/53). ChatGPT□4.0 delivered slightly lower performance in corneal diseases at 85% (17/20) but showed an improved accuracy in glaucoma at 63.6% (7/11) while matching DeepSeek’s performance in neuro□ophthalmology at 81.8%, thereby also achieving an overall accuracy of 79.2% (42/53). In contrast, ChatGPT□01 maintained the same corneal and glaucoma accuracies as ChatGPT□4.0 (85% and 63.6%, respectively) but excelled in neuro□ophthalmology with a remarkable 95.5% (21/22), culminating in the highest overall accuracy of 84.9% (45/53). Conversely, Qwens demonstrated lower performance, with only 55% accuracy in corneal diseases (11/20), 54.5% in glaucoma (6/11), and 77.3% in neuro□ophthalmology, which translated to an overall accuracy of 64.2% (34/53). These results highlight significant variability in diagnostic performance across different AI models and ophthalmologic subspecialties.

Inter□observer agreement with the average human expert performance—defined as 93.3% for corneal diseases (attendings: 100%, 90%, and 90%), 51.5% for glaucoma (attendings: 6/11, 6/11, and 5/11), and 90.9% for neuro□ophthalmology (20/22 for both attendings)—was assessed for each AI model. In corneal diseases, DeepSeek’s accuracy was within 3.3 percentage points of the human average (90% vs 93.3%), whereas both ChatGPT□4.0 and ChatGPT□01 were 8.3 percentage points lower (85% vs 93.3%), and Qwens was 38.3 percentage points lower (55% vs 93.3%). In glaucoma, DeepSeek’s performance exceeded the human average by 3.0 percentage points (54.5% vs 51.5%), while ChatGPT□4.0 and ChatGPT□01 surpassed the human benchmark by 12.1 percentage points (63.6% vs 51.5%); Qwens’ performance was 3.0 percentage points above the human average (54.5% vs 51.5%). In neuro□ophthalmology, both DeepSeek and ChatGPT□4.0 were 9.1 percentage points lower than the human average (81.8% vs 90.9%), whereas ChatGPT□01 outperformed the human experts by 4.6 percentage points (95.5% vs 90.9%), and Qwens trailed by 13.6 percentage points (77.3% vs 90.9%).

**Table 1.** Diagnostic Performance of AI Models in Three Ophthalmologic Specialties.

| AI Model | Specialty | Cases Correct/Total | Percent Correct | Overall Accuracy |
| --- | --- | --- | --- | --- |
| DeepSeek | Corneal diseases | 18 of 20 | 90.0% | 42 of 53 (79.2%) |
|  | Glaucoma | 6 of 11 | 54.5% |  |
|  | Neuro-ophthalmology | 18 of 22 | 81.8% |  |
| **ChatGPT-4.0** | Corneal diseases | 17 of 20 | 85.0% | 42 of 53<br>(79.2%) |
|  | Glaucoma | 7 of 11 | 63.6% |  |
|  | Neuro-ophthalmology | 18 of 22 | 81.8% |  |
| **ChatGPT-01** | Corneal diseases | 17 of 20 | 85.0% | 45 of 53<br>(84.9%) |
|  | Glaucoma | 7 of 11 | 63.6% |  |
|  | Neuro-ophthalmology | 21 of 22 | 95.5% |  |
| **Qwens** | Corneal diseases | 11 of 20 | 55.0% | 34 of 53<br>(64.2%) |
|  | Glaucoma | 6 of 11 | 54.5% |  |
|  | Neuro-ophthalmology | 17 of 22 | 77.3% |  |

Footnote: Overall accuracy is calculated across all 53 cases.

**Table 2.**
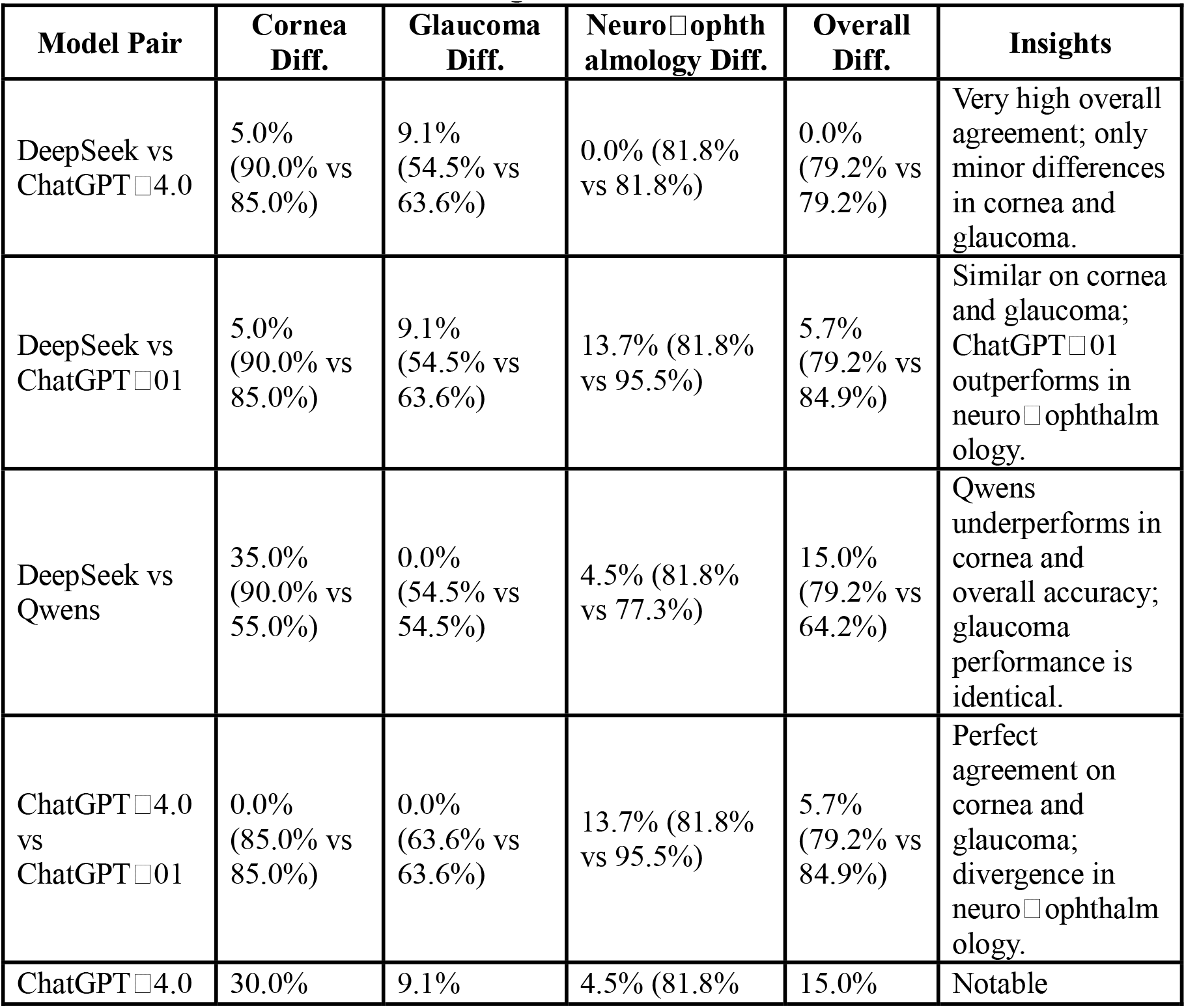
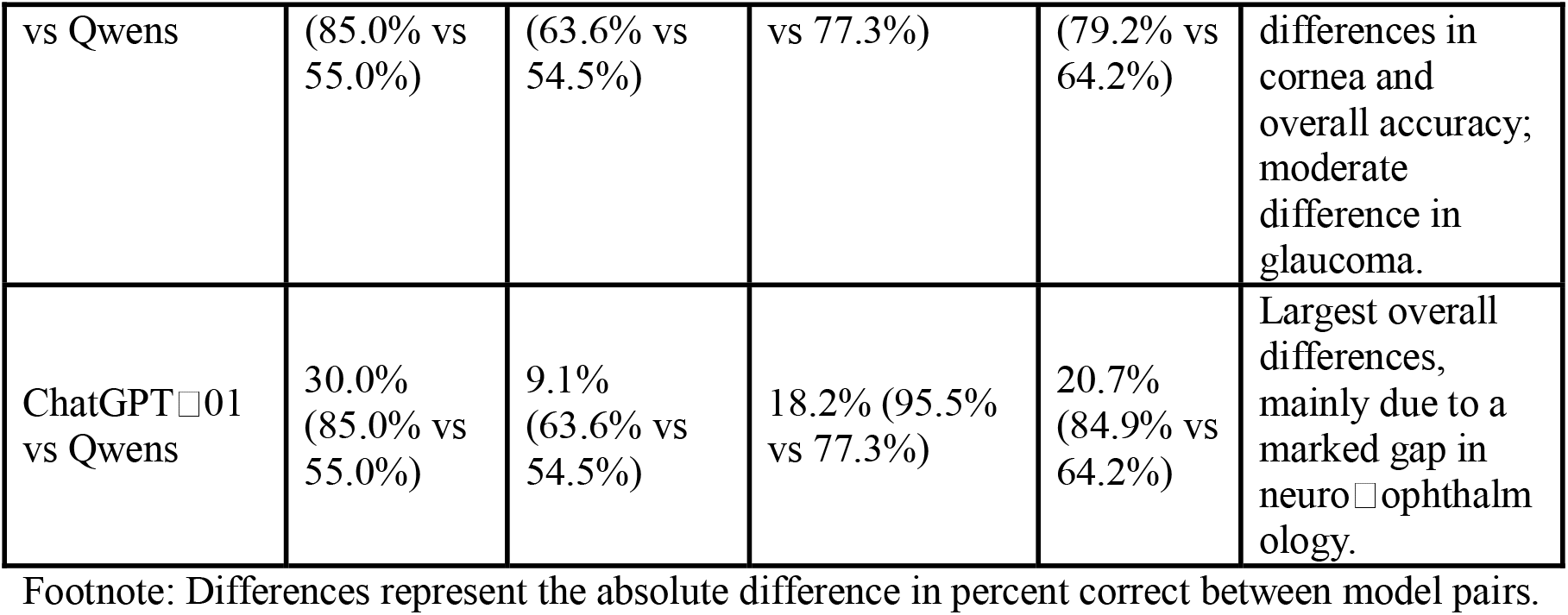
Model Pair Differences in Diagnostic Performance.

**Table 3.** Inter□Observer Agreement for Corneal Diseases.

| AI Model | Human Expert (Avg 93.3%) |
| --- | --- |
| DeepSeek | −3.3% (90% vs 93.3%) |
| ChatGPT-4.0 | −8.3% (85% vs 93.3%) |
| ChatGPT-01 | −8.3% (85% vs 93.3%) |
| Qwens | −38.3% (55% vs 93.3%) |

Footnote: Human expert average derived from three attendings (100%, 90%, and 90%).

**Table 4.** Inter□Observer Agreement for Glaucoma.

| AI Model | Human Expert (Avg 51.5%) |
| --- | --- |
| DeepSeek | +3.0% (54.5% vs 51.5%) |
| ChatGPT-4.0 | +12.1% (63.6% vs 51.5%) |
| ChatGPT-01 | +12.1% (63.6% vs 51.5%) |
| Qwens | +3.0% (54.5% vs 51.5%) |

Footnote: Human expert average derived from three attendings (6/11, 6/11, and 5/11).

**Table 5.**
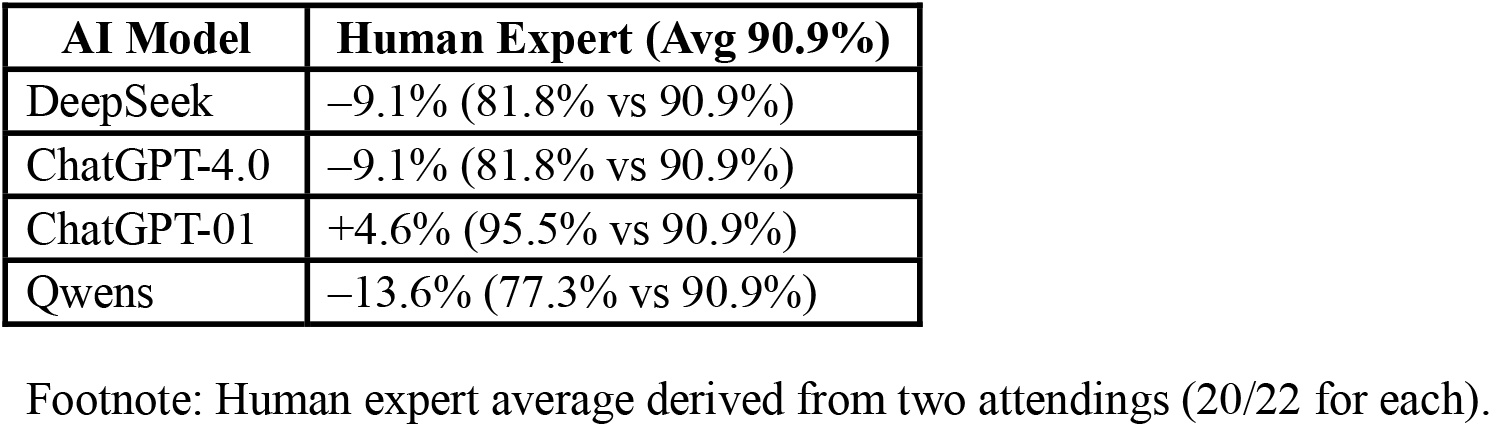
Inter□Observer Agreement for Neuro□Ophthalmology.

These tables succinctly capture the differences in performance between each AI model and the human expert average, with the values representing the percent difference (inter□observer agreement) along with the respective accuracies.

## Discussion

Our findings demonstrate that contemporary large language models exhibit promising yet variable diagnostic capabilities across ophthalmologic subspecialties. Notably, ChatGPT□01 achieved the highest overall accuracy (84.9%), driven by its robust performance in neuro□ophthalmology (95.5% accuracy), where it even surpassed the average human expert performance by 4.6 percentage points. In contrast, both DeepSeek and ChatGPT□4.0 achieved an overall accuracy of 79.2%, with DeepSeek performing comparably to human experts in glaucoma (54.5% vs 51.5%) and only slightly lower in corneal diseases (90.0% vs 93.3%), while ChatGPT□4.0’s performance in glaucoma exceeded the human benchmark by 12.1 percentage points despite similar shortcomings in corneal evaluations. Conversely, Qwens underperformed across the board, particularly in corneal diseases where its accuracy (55.0%) was 38.3 percentage points below that of human experts. These results underscore the potential of AI as an adjunct diagnostic tool in ophthalmology, while also highlighting the need for further refinement, especially in domains such as corneal pathology, to ensure consistent and reliable performance relative to clinical expertise.

In neuro-ophthalmologic cases, our study demonstrated that ChatGPT□01 achieved the highest diagnostic accuracy at 95.5% (21/22), outperforming DeepSeek and ChatGPT□4.0, both at 81.8%, and Qwens at 77.3%. This suggests that ChatGPT□01’s advanced reasoning capabilities may better handle the complex language and intricate details characteristic of neuro□ophthalmology. In comparison, Delsoz et al.’s study reported that GPT□3.5 and GPT□4 diagnosed 59% (13/22) and 82% (18/22) of cases, respectively, with GPT□3.5 showing 55%– 59% agreement with neuro□ophthalmologists and GPT□4 achieving 73%–77% concordance— closely aligning with the experts’ 77% agreement. Notably, our findings diverge from other reports in which GPT□3.5 and GPT□4 demonstrated lower accuracy (40%–48%) on neuro□ophthalmology-specific questions, a discrepancy likely attributable to differences in study methodologies. Moreover, the observed variability between neuro□ophthalmologists in Delsoz et al.’s study underscores the inherent complexity of these diagnoses. Despite these challenges, our results support the potential of AI to serve as a reliable second opinion, complementing human expertise in neuro□ophthalmologic diagnostics.

For corneal diseases, DeepSeek outperformed other models with a diagnostic accuracy of 90.0% (18/20), while Qwens lagged significantly at 55.0% (11/20). ChatGPT-4.0 and ChatGPT-01 delivered intermediate results, both achieving 85.0% (17/20). The nearly 35-percentage point difference between DeepSeek and Qwens suggests substantial differences in how these models interpret corneal pathology, emphasizing the need to understand model-specific training data and algorithmic biases, particularly in anterior segment diagnoses. These findings suggest that certain AI models may be more suitable for diagnosing specific conditions based on their underlying training and algorithms, which can significantly impact diagnostic performance. When comparing our results with previous research, including Delsoz et al., we found similar trends. In their study, ChatGPT-4 made the correct diagnosis in 17 out of 20 cases (85%), while ChatGPT-3.5 showed a lower accuracy of 12 out of 20 cases (60%). In contrast, three cornea specialists achieved diagnostic accuracy between 90% and 100%, with one reaching a perfect score (20/20). This comparison demonstrates that ChatGPT-4’s performance in our study aligns closely with previous research (85% accuracy). However, the marked performance difference between ChatGPT-3.5 (60%) and ChatGPT-4.0 (85%) mirrors earlier findings of significant improvements between GPT versions. These results suggest that newer GPT models may be better suited to specific domains like corneal diseases, but clinicians should exercise caution due to performance variability.

Glaucoma diagnosis remains a persistent challenge for all models, with accuracies ranging narrowly between 54.5% (observed in DeepSeek and Qwens with 6/11 correct) and 63.6% (achieved by ChatGPT-4.0 and ChatGPT-01 with 7/11 correct). These modest performance rates mirror the diagnostic difficulties faced even by human clinicians and underscore the inherent challenges of relying solely on clinical case reports, where subtle clinical signs and nuanced patient history are critical for accurate diagnosis. While prior studies reported a slightly higher accuracy for ChatGPT-3.0 (approximately 72.7%, diagnosing 8/11 cases) and ophthalmology residents demonstrated variability between 54.5% and 72.7% correct diagnoses, the overall agreement between ChatGPT-3.0 and residents was relatively high. Common glaucoma cases were reliably identified (with ChatGPT correctly diagnosing all 8 common cases). However, our models struggled with atypical presentations; this aligns with our earlier work showing that ChatGPT struggled with uncommon glaucoma phenotypes (e.g., aqueous misdirection, inflammatory glaucoma)^18^, where clinical ambiguity and overlapping symptomatology demand integration of imaging and longitudinal data. These results emphasize that while LLMs excel at text-based reasoning, their utility in glaucoma may remain limited until they can process multimodal inputs.

Despite these subspecialty disparities, DeepSeek and ChatGPT-4.0 achieved identical overall accuracy (79.2%), masking their domain-specific strengths and weaknesses. For example, DeepSeek outperformed ChatGPT-4.0 in corneal cases (90.0% vs. 85.0%), while ChatGPT-4.0 showed marginally better performance in glaucoma (63.6% vs. 54.5%). This divergence highlights the inadequacy of aggregated metrics for evaluating clinical AI tools, as similar overall performance can obscure critical differences in subspecialty accuracy differences that may determine whether a model is suitable for triage in a general clinic versus a referral center. Furthermore, the variability in overall accuracy across models (64.2% for Qwens to 84.9% for ChatGPT-01) underscores the need for rigorous, condition-specific validation. A 20-percentage-point gap in accuracy could translate to significant clinical consequences, such as delayed referrals or mismanagement of sight-threatening conditions like neuro-ophthalmic emergencies.

These findings also raise questions about complementary model integration. For instance, ChatGPT-01’s neuro-ophthalmology expertise could compensate for Qwens’ weaknesses in corneal diseases, suggesting that ensemble approaches rather than reliance on a single model might offer more robust diagnostic support. This strategy aligns with emerging trends in AI, where hybrid systems combine multiple models to mitigate individual limitations^19,20^. However, such approaches require careful calibration to avoid overcomplicating clinical workflows or introducing conflicting recommendations.

The study’s limitations must also be acknowledged. First, the small sample size limits generalizability. Larger, multicenter datasets are needed to validate these findings, though ethical and logistical challenges in curating such datasets remain significant. Second, the exclusion of imaging restricts the models’ diagnostic potential, especially for glaucoma and neuro-ophthalmology, where fundus photos, OCT scans, and visual fields are integral to decision-making. However, we did include the interpretation of imaging findings in relevant cases. Future iterations of LLMs that integrate multimodal inputs could bridge this gap, as seen in preliminary studies combining text and imaging in AI diagnostics^21^. Third, the risk of hallucinations and factually incorrect outputs remains a concern, particularly for rare conditions underrepresented in training data (e.g., lattice corneal dystrophy)^12,22^. Clinicians must therefore treat LLM-generated diagnoses as provisional, requiring validation against expert judgment.

## Data Availability

All data produced in the present work are contained in the manuscript.

## Acknowledgements

None.

## Funding

This work was supported by NIH Grants R01EY033005 (SY) and supports from the Hamilton Eye Institute (SY). The funders had no role in study design, data collection and analysis, decision to publish, or preparation of the manuscript.

## Conflicts of interest

Zain S. Hussain, Mohammad Delsoz, Muhammad Elahi, Brian Jerkins, Elliot Kanner, Claire Wright, Wuqaas M. Munir, Mohammad Soleimani, Ali Djalilian, Priscilla A. Lao, Joseph W. Fong, and Malik Y. Kahook have nothing to disclose. Siamak Yousefi: Received prototype instruments from Remidio, M&S Technologies, and Virtual Fields. He provides consultations to InsightAEye and Enolink.

## Data sharing statement

All essential data for replicating our results is in the Supplementary file, available upon requests.

## Notes

### Competing Interest Statement

The authors have declared no competing interest.

### Funding Statement

This study did not receive any funding.

