## Supplemental Table 1 for "Performance of DeepSeek, Qwen 2.5 MAX, and ChatGPT Assisting in Diagnosis of Corneal Eye Diseases, Glaucoma, and Neuro-Ophthalmology Diseases Based on Clinical Case Reports"

| **Supplementary Table 1.** Cornea - Provisional diagnoses provided by ChatGPT-4.0, ChatGPT-o1, DeepSeek-V3, Qwen-2.5 Max  and Human Experts | | | | | | |
| --- | --- | --- | --- | --- | --- | --- |
| No | Corneal Eye diseases | ChatGPT4 Diagnosis | ChatGPT-o1 | DeepSeek-V3 | Qwen-2.5 Max | Human Expert Diagnosis |
| 1 | Cystinosis | Cystinosis | Cystinosis | Cystinosis | Cystinosis | H1: Cystinosis  H2: Cystinosis  H3: Cystinosis |
| 2 | Fuchs’ Endothelial Corneal Dystrophy (FECD) | FECD | FECD | FECD | Posterior Polymorphous  Corneal Dystrophy (PPCD) | H1: FECD  H2: FECD  H3: FECD |
| 3 | Pseudophakic Bullous Keratopathy (PBK) | PBK | PBK | PBK | Corneal decompensation secondary to endothelial dysfunction | H1: PBK  H2: PBK  H3: PBK |
| 4 | Amiodarone-Induced Corneal Deposits (Corneal Verticillata) | Amiodarone-Induced Corneal Deposits | Amiodarone-Induced Corneal Deposits | Amiodarone-Induced Corneal Deposits | Amiodarone-Induced Corneal Deposits | H1: Amiodarone-Induced Corneal Deposits  H2: Amiodarone-Induced Corneal Deposits  H3: Amiodarone-Induced Corneal Deposits |
| 5 | Acanthamoeba Keratitis | Acanthamoeba Keratitis | s, Acanthamoeba keratitis (stage 5) | Acanthamoeba Keratitis | Acanthamoeba Keratitis | H1: Acanthamoeba Keratitis  H2: Acanthamoeba Keratitis  H3: Acanthamoeba Keratitis |
| 6 | Cogan's Syndrome (Interstitial Keratitis & Vertigo) | Scleritis | Cogan’s syndrome | anterior scleritis | Scleritis | H1: Cogan's Syndrome  H2: Episcleritis  H3: Cogan's Syndrome |
| 7 | Infectious Crystalline Keratopathy (ICK) | Fungal Keratitis | ICK | ICK | Recurrent Simplex Viral Keratitis | H1: ICK  H2: ICK  H3: ICK |
| 8 | Megalocornea | Megalocornea | subluxation (instability) of the iris‐sutured intraocular lens. | Megalocornea | pseudophakic pupillary block glaucoma | H1: Megalocornea  H2: Megalocornea  H3: Megalocornea |
| 9 | Herpes Simplex Viral Keratitis | Herpes Simplex Viral Keratitis | Herpes simplex keratitis | Herpes Simplex Viral Keratitis | Herpes Simplex Viral Keratitis | H1: Herpes Simplex Viral Keratitis  H2: Herpes Simplex Viral Keratitis  H3: Herpes Simplex Viral Keratitis |
| 10 | Atopic Keratoconjunctivitis | Atopic Keratoconjunctivitis |  | Atopic Keratoconjunctivitis | Ocular Cicatricial Pemphigoid (OCP) | H1: Atopic Keratoconjunctivitis  H2: Atopic Keratoconjunctivitis  H3: OCP |
| 11 | Lattice Corneal Dystrophy Type II (Meretoja’s syndrome) | Meesmann Corneal Dystrophy (MCD) | Lattice Corneal Dystrophy Type II (Meretoja syndrome). | Lattice Corneal Dystrophy I | Meesmann Corneal Dystrophy | H1: Lattice Corneal Dystrophy Type II  H2: Lattice Corneal Dystrophy Type II  H3: Lattice Corneal Dystrophy Type II |
| 12 | Salzmann's Nodular Degeneration (SND) | SND | Salzmann's nodular degeneration | granular corneal dystrophy type 2 (Avellino corneal dystrophy) | SND | H1: SND  H2: SND  H3: SND |
| 13 | Exposure Keratopathy | Exposure Keratopathy | Exposure Keratopathy | Exposure Keratopathy | Bilateral bacterial keratitis with corneal epethelial defects and dellen formation | H1: Exposure Keratopathy  H2: Exposure Keratopathy  H3: Exposure Keratopathy |
| 14 | Peripheral Ulcerative Keratitis | Peripheral Ulcerative Keratitis | peripheral ulcerative keratitis (PUK) associated with rheumatoid arthritis. | Peripheral Ulcerative Keratitis | Peripheral Ulcerative Keratitis | H1: Peripheral Ulcerative Keratitis  H2: Peripheral Ulcerative Keratitis  H3: Peripheral Ulcerative Keratitis |
| 15 | Calcific Band Keratopathy | Calcific Band Keratopathy | Calcific band-shaped keratopathy | Calcific Band Keratopathy | Calcific Band Keratopathy | H1: Calcific Band Keratopathy  H2: Calcific Band Keratopathy  H3: Calcific Band Keratopathy |
| 16 | Posterior Polymorphous  Corneal Dystrophy (PPCD) | PPCD | fleck corneal dystrophy | posterior amorphous corneal dystrophy (PACD) | posterior amorphous corneal dystrophy (PACD) | H1: PPCD  H2: Granular Corneal Dystrophy  H3: PPCD |
| 17 | Acute Corneal Hydrops | Acute Corneal Hydrops | acute corneal hydrops | Acute Corneal Hydrops | Acute Corneal Hydrops | H1: Acute Corneal Hydrops  H2: Acute Corneal Hydrops  H3: Acute Corneal Hydrops |
| 18 | Corneal Marginal  Ulcer | Corneal Ulceration | staphylococcal marginal keratitis | Corneal Ulcer | Corneal ulcer associated with keratitis | H1: Corneal Marginal Ulcer  H2: Corneal Marginal Ulcer  H3: Mooren Ulcer |
| 19 | Fabry Disease | Fabry Disease | Fabry disease | Fabry Disease | Fabry Disease | H1: Fabry Disease  H2: Fabry Disease  H3: Fabry Disease |
| 20 | Cytarabine Induced Keratoconjunctivitis | Cytarabine Induced Keratoconjunctivitis | cytarabine-induced keratoconjunctivitis | Cytarabine Induced Keratoconjunctivitis | Cytarabine Induced Keratoconjunctivitis | H1: Cytarabine Induced Keratoconjunctivitis  H2: Cytarabine Induced Keratoconjunctivitis  H3: Cytarabine Induced Keratoconjunctivitis |
