## Supplemental Table 2 for "Performance of DeepSeek, Qwen 2.5 MAX, and ChatGPT Assisting in Diagnosis of Corneal Eye Diseases, Glaucoma, and Neuro-Ophthalmology Diseases Based on Clinical Case Reports"

| **Supplementary Table 2.** Glaucoma - Provisional diagnoses provided by ChatGPT-4.0, ChatGPT-o1, DeepSeek-V3, Qwen-2.5 Max  and Human Experts | | | | | | | | |
| --- | --- | --- | --- | --- | --- | --- | --- | --- |
| **Cases** | **Name of disease** | **DeepSeek diagnosis** | **ChatGPT4 Diagnosis** | **ChatGPT-o1 diagnosis** | **Qwen-2.5 Max Diagnosis** | **Glaucoma Attending 1** | **Glaucoma Attending 2** | **Glaucoma Attending 3** |
| **Primary open-angle glaucoma (POAG)** | Juvenile open-angle glaucoma | Juvenile open-angle glaucoma | Juvenile open-angle glaucoma | Juvenile open-angle glaucoma | Juvenile open-angle glaucoma | Juvenile open-angle glaucoma | Juvenile open-angle glaucoma | Juvenile open-angle glaucoma |
|  | Normal Tension Glaucoma | Normal Tension Glaucoma | Normal Tension Glaucoma | Normal Tension Glaucoma | Normal Tension Glaucoma | Low Tension Glaucoma* | Low Tension Glaucoma* | Low Tension Glaucoma* |
|  | refractory primary open angle glaucoma | medically-uncontrolled primary open-angle glaucoma | medically-uncontrolled primary open-angle glaucoma | medically-uncontrolled primary open-angle glaucoma | medically-uncontrolled primary open-angle glaucoma | Uveitic Glaucoma | Uveitic Glaucoma | Uveitic Glaucoma |
| **Secondary open angle glaucoma** | Pseudo-exfoliations glaucoma | Pseudo-exfoliations glaucoma | Pseudo-exfoliations glaucoma | Pseudo-exfoliations glaucoma | Pseudo-exfoliations glaucoma | Pseudo-exfoliations glaucoma | Pseudo-exfoliations glaucoma | Pseudo-exfoliations glaucoma |
|  | pigmentary dispersion syndrome | pigmentary dispersion syndrome | pigmentary dispersion syndrome | pigmentary dispersion syndrome | pigmentary dispersion syndrome | pigmentary dispersion syndrome | pigmentary dispersion syndrome | pigmentary dispersion syndrome |
|  | Glaucomatocyclitic Crisis | acute angle-closure glaucoma | Glaucomatocyclitic Crisis | Recurrent uveitic glaucoma | recurrent secondary angle-closure glaucoma | Glaucomatocyclitic Crisis | Fuch's heterochromic iridocyclitis | CACG with acute IOP rise phase |
|  | Aphakic glaucoma | Aphakic glaucoma | Aphakic glaucoma | Aphakic glaucoma | Aphakic glaucoma | Aphakic Glaucoma | Aphakic Glaucoma | Aphakic Glaucoma |
| **Primary angle closure glaucoma** | congenital glaucoma | congenital glaucoma | congenital glaucoma | congenital glaucoma | congenital glaucoma | Congenital Glaucoma | Congenital Glaucoma | Congenital Glaucoma |
| **Secondary angle closure glaucoma** | Neovascular glaucoma | central retinal vein occlusion | central retinal vein occlusion | central retinal vein occlusion | central retinal vein occlusion | central retinal vein occlusion | central retinal vein occlusion | central retinal vein occlusion |
|  | aqueous misdirection | acute angle-closure glaucoma | acute angle-closure glaucoma | aqueous misdirection/malignant glaucoma | acute angle-closure glaucoma | Aqueous Misdirection | Aqueous Misdirection | Aqueous misdirection |
|  | Grant syndrome | primary angle-closure glaucoma | Glaucomatocyclitic Crisis | Acute angle closure glaucoma | Chronic Angle Closure Glaucoma | Chronic Angle Closure Glaucoma | Chronic Acute-Angle Closure Glaucoma | Fuch's heterochromic iridocyclitis |
| **Total cases =11** |  |  |  |  |  |  |  |  |

Table Footenotes: Normal Tension Glaucoma is same disease entity as Low Tension Glaucoma
