## Supplemental Table 3 for "Performance of DeepSeek, Qwen 2.5 MAX, and ChatGPT Assisting in Diagnosis of Corneal Eye Diseases, Glaucoma, and Neuro-Ophthalmology Diseases Based on Clinical Case Reports"

| **Supplementary Table 3.** Neuro-ophthalmology - Provisional diagnoses provided by ChatGPT-4.0, ChatGPT-o1, DeepSeek-V3, Qwen-2.5 Max  and Human Experts | | | | | | |
| --- | --- | --- | --- | --- | --- | --- |
| Correct diagnosis of the case | deepseek diagnosis | GPT-4 diagnosis | GPT-o1 diagnosis | Qwen 2.5 diagnosis | Neuro-ophthalmologist 1 diagnosis | Neuro-ophthalmologist 2 diagnosis |
| Demyelinating Encephalomyelitis (ADEM) with Associated Optic Neuritis | MOG antibody-associated disease (MOGAD) | Neuromyelitis Optica Spectrum Disorder (NMOSD) | Disseminated Encephalomyelitis (ADEM) | Disseminated Encephalomyelitis (ADEM) | Optic Neuritis 2/2 Meningitis | Viral Meningitis |
| Optic Nerve Drusen | optic disc drusen | Optic Disc Drusen | bilateral optic disc drusen | Optic Disc Drusen | Optic Disc Drusen | Optic Disc Drusen |
| Optic Nerve Hypoplasia | optic nerve hypoplasia | Mature Cataract and Optic Atrophy in the Left Eye, along with Exotropia | optic nerve hypoplasia | chronic optic neuropathy | Optic Nerve Hypoplasia | Optic Nerve Hypoplasia |
| Optic Neuritis | optic neuritis as part of multiple sclerosis. | Optic Neuritis | Optic Neuritis 2/2 MS | Optic Neuritis 2/2 MS | Optic Neuritis 2/2 MS | Multiple Sclerosis, Optic Neuritis |
| Optic Nerve Hypoplasia | Septo-optic Dysplasia | Optic Nerve Hypoplasia (ONH) | septo-optic dysplasia (de Morsier syndrome) | septo-optic dysplasia (de Morsier syndrome) | Demorsier Syndrome | Septo-Optic Dysplasia with Optic Nerve Hypoplasia |
| Cranial Nerve IV (Trochlear Nerve) Palsy | right fourth cranial nerve (trochlear nerve) palsy | Fourth Nerve Palsy (Trochlear Nerve Palsy) | right fourth cranial nerve (trochlear nerve) palsy. | right fourth cranial nerve (trochlear nerve) palsy. | Traumatic CN IV Palsy OD | Right Traumatic Fourth Nerve Palsy |
| Dorsal Midbrain Syndrome (Parinaud's Syndrome) | ocular tilt reaction (OTR) secondary to a brainstem hemorrhage involving the midbrain and upper pons. | Parinaud's Syndrome, (Dorsal Midbrain Syndrome) | dorsal midbrain syndrome (also known as Parinaud’s syndrome) | Dorsal Midbrain Syndrome | Dorsal Midbrain Syndrome | Dorsal Midbrain Syndrome |
| Traumatic Optic Neuropathy | traumatic optic neuropathy (TON) | Traumatic Optic Neuropathy (TON) | traumatic optic neuropathy | Traumatic Optic Neuropathy (TON) | Traumatic Optic Neuropathy 2/2 Orbital Canal Fracture | Traumatic Optic Neuropathy |
| Ethambutol Toxicity and Optic Neuropathy | Ethambutol-Induced Optic Neuropathy | Toxic Optic Neuropathy, most likely related to the use of Ethambutol | ethambutol-induced optic neuropathy | ethambutol-induced optic neuropathy | Ethambutol Toxicity/Toxic Optic Neuropathy | Toxic (Ethambutol) Optic Neuropathy |
| Posterior Ischemic Optic Neuropathy | Posterior Ischemic Optic Neuropathy (PION) | Perioperative Ischemic Optic Neuropathy (PION) | bilateral posterior ischemic optic neuropathy (PION) | bilateral posterior ischemic optic neuropathy (PION) | PION | Posterior Ischemic Optic Neuropathy, Bilateral |
| Recurrent Neuroretintitis | Multiple Sclerosis (MS) | Optic Neuritis (ON) | neuroretinitis | Optic Neuritis | Optic Neuritis | Optic Neuritis |
| Horner's Syndrome due to Cluster Headache | Cluster Headache, specifically the variant known as "Cluster-Tic Syndrome" or "Cluster Migraine Syndrome | Cluster Headache (CH), specifically with a Horner's syndrome | cluster headaches | Cluster Headache (CH), specifically with a Horner's syndrome | Horner Syndrome 2/2 Cluster Headaches | Cluster Headache |
| Horner Syndrome due to Ipsilateral Internal Carotid Artery Dissection | Carotid Artery Dissection | Horner's Syndrome related to a Carotid Artery Dissection | Horner Syndrome 2/2 Carotid Dissection | Horner Syndrome 2/2 Carotid Dissection | Horner Syndrome 2/2 Carotid Dissection | Cervical Carotid Artery Dissection Causing Horner Syndrome |
| Idiopathic Intracranial Hypertension (Pseudotumor Cerebri) | Idiopathic Intracranial Hypertension (IIH), also known as Pseudotumor Cerebri | Idiopathic Intracranial Hypertension (IIH), also known as Pseudotumor Cerebri | Idiopathic Intracranial Hypertension (IIH) | Idiopathic Intracranial Hypertension (IIH) | IIH | Idiopathic Intracranial Hypertension |
| Idiopathic Orbital Myositis | orbital myositis, a subtype of idiopathic orbital inflammatory syndrome (IOIS) | Idiopathic Orbital Inflammatory Syndrome (IOIS), also known as Orbital Myositis | idiopathic orbital myositis | Idiopathic Orbital Inflammatory Syndrome (IOIS), also known as Orbital Myositis | Orbital Myositis | Thyroid Eye Disease |
| Leber Hereditary Optic Neuropathy | Leber Hereditary Optic Neuropathy | Papilledema | Leber Hereditary Optic Neuropathy | toxic-nutritional optic neuropathy | LHON | Leber’s Hereditary Optic Neuropathy |
| Miller Fisher Syndrome | Miller Fisher Syndrome | Miller Fisher Syndrome (MFS) | acute ophthalmoplegia variant of Guillain-Barré syndrome (on the Miller Fisher spectrum). | Miller Fisher Syndrome | Miller Fisher/Guillan Barre | Miller Fisher Syndrome |
| Myasthenia Gravis | unilateral congenital ptosis with possible Marcus Gunn jaw-winking syndrome. | Myasthenia Gravis (MG) | ocular myasthenia gravis | congenital ptosis with poor levator function . | Myasthenia Gravis | Ocular Myasthenic Gravis |
| Neurofibromatosis Type 1—Optic Nerve Glioma | Optic Pathway Glioma | Neurofibromatosis Type 1 (NF1) | optic pathway glioma (optic nerve glioma) associated with NF1 | Neurofibromatosis Type 1—Optic Nerve Glioma | Optic Nerve Glioma | Optic Pathway Glioma |
| Thyroid Eye Disease (Graves' Ophthalmopathy) | Thyroid Eye Disease (Graves' Ophthalmopathy) | Thyroid Eye Disease (TED), also known as Graves' Ophthalmopathy | thyroid eye disease (Graves’ orbitopathy) | Thyroid Eye Disease (Graves' Ophthalmopathy) | TED | Thyroid Eye Disease with Compressive Optic Neuropathy |
| Visual Snow Syndrome | Visual Snow Syndrome | Visual Snow Syndrome (VSS) | Visual Snow Syndrome | Visual Snow Syndrome | Visual Snow | Visual Snow Syndrome |
| Unilateral Optic Nerve Hypoplasia | Congenital Fibrosis of the Extraocular Muscles (CFEOM), specifically CFEOM type 1 | Optic Nerve Hypoplasia (ONH) | left optic nerve hypoplasia | Unilateral Optic Nerve Hypoplasia | Sensory Exotropia 2/2 Optic Nerve Hypoplasia | Optic Nerve Hypoplasia |
